# A genetically based computational drug repurposing framework for rapid identification of candidate compounds: application to COVID-19

**DOI:** 10.1101/2025.01.10.25320348

**Authors:** Georgios Voloudakis, Kyung Min Lee, James M. Vicari, Wen Zhang, Daisy Hoagland, Sanan Venkatesh, Jiantao Bian, Marios Anyfantakis, Zhenyi Wu, Samir Rahman, Lina Gao, Kelly Cho, Jennifer S. Lee, Sudha K. Iyengar, Shiuh-Wen Luoh, Themistocles L. Assimes, Gabriel E. Hoffman, Benjamin R. tenOever, John F. Fullard, Julie A. Lynch, Panos Roussos

**Affiliations:** Center for Disease Neurogenomics, Icahn School of Medicine at Mount Sinai, New York, NY USA; Department of Psychiatry, Icahn School of Medicine at Mount Sinai, New York, NY, USA; Friedman Brain Institute, Icahn School of Medicine at Mount Sinai, New York, NY, USA; Department of Genetics and Genomic Science, Icahn School of Medicine at Mount Sinai, New York, NY, USA; Department of Artificial Intelligence and Human Health, Icahn School of Medicine at Mount Sinai, New York, NY, USA; Mental Illness Research, Education, and Clinical Center (VISN 2 South), James J. Peters VA Medical Center, Bronx, NY, USA; Center for Precision Medicine and Translational Therapeutics, James J. Peters VA Medical Center, Bronx, NY, USA; VA Informatics and Computing Infrastructure, VA Salt Lake City Health Care System, Salt Lake City, UT, USA; Department of Microbiology, Icahn School of Medicine at Mount Sinai, New York, NY, USA; Virus Engineering Center for Therapeutics and Research, Icahn School of Medicine at Mount Sinai, New York, NY, USA; Global Health and Emerging Pathogens Institute, Icahn School of Medicine at Mount Sinai, New York, NY, USA; Division of Epidemiology, University of Utah, Salt Lake City, UT, USA; Biostatistics Shared Resources, Knight Cancer Institute, Oregon Health & Science University, Portland, OR, USA; VA Boston Healthcare System, Boston, MA, USA; Division of Aging, Brigham and Women’s Hospital, Harvard Medical School, Boston, MA, USA; Department of Medicine, Stanford University School of Medicine, Stanford, CA, USA; VA Palo Alto Health Care System, Palo Alto, CA, USA; Department of Population and Quantitative Health Sciences, School of Medicine, Case Western Reserve University, Cleveland, OH, USA; Department of Genetics and Genome Sciences, School of Medicine, Case Western Reserve University, Cleveland, OH, USA; VA Northeast Ohio Healthcare System, Cleveland VA Medical Center, Cleveland, OH, USA; Department of Medicine, Knight Cancer Institute, Oregon Health & Science University, Portland, OR, USA; VA Portland Health Care System, Portland, OR, USA

**Keywords:** TWAS, computational drug repurposing, pharmacoepidemiology, COVID-19

## Abstract

**Background:** The development and approval of novel drugs are typically time-intensive and expensive. Leveraging a computational drug repurposing framework that integrates disease-relevant genetically regulated gene expression (GReX) and large longitudinal electronic medical record (EMR) databases can expedite the repositioning of existing medications. However, validating computational predictions of the drug repurposing framework remains a challenge.

**Methods:** To benchmark the drug repurposing framework, we first performed a 5-method-rank-based computational drug prioritization pipeline by integrating multi-tissue GReX associated with COVID-19-related hospitalization, with drug transcriptional signature libraries from the Library of Integrated Network-Based Cellular Signatures. We prioritized FDA-approved medications from the 10 top-ranked compounds, and assessed their association with COVID-19 incidence within the Veterans Health Administration (VHA) cohort (~9 million individuals). In parallel, we evaluated *in vitro* SARS-CoV-2 replication inhibition in human lung epithelial cells for the selected candidates.

**Results:** Our *in silico* pipeline identified seven FDA-approved drugs among the top ten candidates. Six (imiquimod, nelfinavir and saquinavir, everolimus, azathioprine, and retinol) had sufficient prescribing rates or feasibility for further testing. In the VHA cohort, azathioprine (odds ratio [OR]=0.69, 95% CI 0.62–0.77) and retinol (OR=0.81, 95% CI 0.72–0.92) were significantly associated with reduced COVID-19 incidence. Conversely, nelfinavir and saquinavir demonstrated potent SARS-CoV-2 inhibition *in vitro* (~95% and ~65% viral load reduction, respectively). No single compound showed robust protection in both *in vivo* and *in vitro* settings.

**Conclusions:** These findings underscore the power of GReX-based drug repurposing in rapidly identifying existing therapies with potential clinical relevance; four out of six compounds showed a protective effect in one of the two validation approaches. Crucially, our results highlight how a complementary evaluation—combining epidemiological data and *in vitro* assays—helps refine the most promising candidates for subsequent mechanistic studies and clinical trials. This integrated validation approach may prove vital for accelerating therapeutic development against current and future health challenges.

## Background

The traditional process of developing and approving novel therapeutics is notoriously time-consuming and expensive, often requiring more than a decade and substantial financial resources, averaging over $2.8 billion per drug [1]. This lengthy process poses significant challenges, especially in the face of emerging health threats or diseases lacking effective treatments. Drug repurposing—the practice of identifying new therapeutic uses for existing medications—offers a promising alternative that circumvents many of the barriers associated with *de novo* drug development. By leveraging known safety profiles and existing clinical data, drug repurposing provides a cost-effective and rapid pathway to bring medications into clinical practice or guide a second wave of targeted drug development efforts.

Despite its promise, the success of drug repurposing efforts hinges on the availability of robust and scalable methodologies to identify and validate candidate compounds. Advances in computational approaches [2], coupled with the growing availability of electronic medical records (EMRs) [3] and *in vitro* validation platforms [4, 5], have revolutionized the landscape of drug repurposing. Integrating genetically regulated gene expression (GReX) data with perturbagen signature libraries, for instance, allows for the identification of compounds capable of reversing disease-associated predicted gene expression dysregulation [6, 7]. However, a critical gap remains in systematically validating these computational predictions to ensure their translational potential.

To address this need, we developed a comprehensive computational drug repurposing framework that integrates multi-tissue GReX with EMR-based pharmacoepidemiological analysis and *in vitro* assays. This pipeline not only prioritizes candidate compounds but also provides a benchmark for their validation using orthogonal approaches. By combining computational predictions with real-world evidence from EMR data and mechanistic insights from *in vitro* experiments, our framework offers a robust, scalable solution for accelerating drug discovery and development.

As a proof of concept, we applied this pipeline to the context of COVID-19, a global health crisis that demanded unprecedented speed in therapeutic discovery. This application demonstrates the versatility and efficacy of our approach, highlighting its potential to inform drug repurposing efforts across a wide range of diseases and clinical scenarios.

## Methods

### GReX-based computational drug repurposing

#### COVID-19 GReX

Publicly available COVID-19 transcriptome-wide association study (TWAS) summary statistics were obtained from our prior study [8]. Specifically, we used the TWAS based on the “hospitalized COVID vs. population” (B2) phenotype from the COVID-19 Host Genetics initiative [9] (Release 4; 2020-10-20; https://www.covid19hg.org/results/r4/). Multi-tissue TWAS input data utilized transcriptomic imputation models from 17 tissues (Table S1; tissues with significant gene-trait associations after FDR [10] adjustment was applied to A1, A2, B1, B2, C1, C2, and D1 COVID-19 phenotypes, and 42 transcriptomic imputation models) trained with EpiXcan [7] on two independent cohorts: GTEx (v8 [11]) and STARNET [12].

#### Perturbagen library

We leveraged the compounds in the LINCS Phase II L1000 dataset (GSE70138) perturbagen reference library [13]. All inferred genes (n=12,328) were considered. Only “gold” (si_gold) signatures were analyzed.

#### Computational drug repurposing (CDR)

For CDR, we integrated GReX with drugs in LINCS. Each signature from the perturbagen signature library (e.g. treatment with x compound for n hours in MCF7 cells) is assessed and ranked for its ability to reverse the trait-associated imputed transcriptomes using a 5-method-rank approach [6] (with 100 permutations). Thus, analysis was limited to 495 potentially repurposable compounds. Drug information for the compounds under consideration (e.g. clinical phase, mechanism of action, and molecular targets) was obtained from http://www.broadinstitute.org/repurposing (file date: 3/24/2020). The compounds under investigation were compared with all the other compounds. For the “mechanism of action comparison”, all compounds with a known mechanism of action represented by two or more candidates were evaluated. Final recommendations are restricted to launched medications, and FDR correction was applied only to launched compounds.

#### Summarization of the effect of signatures across tissues

To summarize the ability of different signatures to antagonize GReX at the compound and mechanism of action level, we leveraged our previously developed [8] non-parametric ranking approach. Briefly, for each treatment condition (signatures), we obtained the average rank from the 5-method-rank. After pooling these signature-level results, we performed a Mann-Whitney U test for each candidate compound against all other signatures to assess whether a candidate’s rankings significantly deviate from the median. For each candidate, we also estimated a GReX antagonism pseudo-z-score, which is defined as the negative Hodges-Lehmann estimator (of the median difference between that specific candidate vs. the other candidates) divided by the standard deviation of the ranks across all perturbations 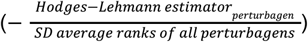. A positive pseudo-z-score suggests that a given compound is a potential therapeutic candidate, whereas a negative pseudo-z-score implies that the compound is likely to exacerbate the phenotype. FDR was estimated using the Benjamini–Hochberg procedure [10]. Signatures were grouped by compound or mechanism of action (minimum requirement of 2 compounds) to derive respective ranks. Signature-GReX combinations of all tissues were considered jointly. Only “launched” medications with 2 or more eligible signatures were retained in the final ranking and FDR [10] correction (Table S2).

#### Software availability

Our R package for computational drug repurposing, antagonist, can be accessed from our center’s GitHub repository: https://github.com/DiseaseNeuroGenomics/antagonist.

### Population-level analysis of the effect of compound and compound category use against COVID-19 incidence

#### Data

We used the VA COVID-19 Shared Data Resource (CSDR), a data domain that includes demographic and clinical information related to COVID-19 on all patients who tested for SARS-CoV-2 within the Veterans Health Administration (VHA) or whose positive test result outside VHA was recorded in VHA clinical notes. The CSDR was supplemented with additional data elements from the VHA’s Corporate Data Warehouse (CDW), a national repository of electronic health records of all individuals who received care in the VHA.

#### Cohort

The base cohort included all Veterans alive as of February 15, 2020. Since the earliest testing date reported in the CSDR was February 16, 2020, we considered all living patients through February 15 to be eligible to be tested for SARS-CoV-2 (Table S3). From this base cohort (Table S4), we derived two separate samples to examine the incidence of COVID-19 among users of the top 10 compounds and antiretroviral medications (Table S5). For the top 10 compound analysis, we assembled a sample of patients who underwent SARS-CoV-2 testing matched to patients who did not undergo SARS-CoV-2 testing on age, race, and VHA facility. The index date was defined as February 15, 2020. For the antiretroviral medication analysis, we created a sample of patients ever diagnosed with human immunodeficiency virus (HIV) prior to the index date and actively on selected antiretroviral medications in the 90 days prior to the index date. Analysis of national VA data was conducted under the protocol, “Leveraging Electronic Health Information to Advance Precision Medicine (LEAP)”, which was approved by the VA Central Institutional Review Board and by the Research & Development Committees at Palo Alto, Salt Lake City, and West Haven VA Medical Centers.

#### Exposure

Exposure to the top 7 FDA-approved compounds (imiquimod, nelfinavir, saquinavir, everolimus, azathioprine, nisoldipine, and retinol), and selected antiretroviral medications (Tables S6 and S7) was determined based on prescription records from the 90 days preceding the index date.

#### Outcome

We used a binary variable indicating a positive reverse transcriptase polymerase chain reaction (RT-PCR) SARS-CoV-2 test result through November 30, 2020.

#### Covariates

We assessed, at the index date, patients’ age, race, marital status, body mass index, smoking status, the Charlson Comorbidity index [14] in the prior two years, VHA utilization in the prior year, the number of days to first SARS-CoV-2 positivity, and the presence of drug-specific FDA-approved and common off-label indications (Table S8) as determined by International Classification of Diseases. We also included the VHA facility of SARS-CoV-2 testing as a fixed effect in the individual compound models and as a random effect in the antiretroviral medication model.

#### Statistical analysis

Multivariable ordinal logistic models were used to test the association of drug exposure with COVID-19 incidence, weighted by the inverse of the predicted probabilities of being tested for SARS-CoV-2 (Tables S9 and S10). Due to the limited availability of SARS-CoV-2 tests and resources, testing was prioritized based on a wide range of factors (e.g., patients’ demographics, comorbidities, and symptom severity). This targeted testing likely resulted in a non-random subset of patients tested for SARS-CoV-2 [15]. To adjust for this, we employed the inverse probability weighting method, where the weight is based on the predicted probabilities (propensity scores) of being tested, estimated by a logistic regression model using drug exposure, selected patient covariates, and VHA facility [16, 17] (Table S11). For this propensity model, we implemented a nested case-control design with incidence density sampling to match each tested patient (case) to five patients who were eligible to be tested (controls) at the time of the case’s testing on age, race, and VHA facility [18].

### In vitro anti-SARS-CoV-2 activity of top candidate compounds

#### Cell line used

Human lung epithelial cells (A549), expressing the angiotensin-converting enzyme 2 receptor (ACE2), the primary entry receptor for SARS-CoV-2, were used for drug treatment and subsequent infection with SARS-CoV-2. These cells, gifted to us, were previously established by the Rosenberg lab to express the ACE2 receptor [4].

#### SARS-CoV-2 virus propagation and infections

SARS-CoV-2, isolate USA-WA1/2020 (NR-52281), was deposited by the Center for Disease Control and Prevention and obtained through BEI Resources, NIAID, NIH. SARS-CoV-2 was propagated in Vero E6 cells in DMEM supplemented with 2% FBS, 4.5 g/L D-glucose, 4 mM L-glutamine, 10 mM Non-Essential Amino Acids, 1 mM Sodium Pyruvate, and 10 mM HEPES. The virus stock was filtered by centrifugation using an Amicon Ultra-15 Centrifugal filter unit (Sigma, Cat # UFC910096) and resuspended in viral propagation media. All infections were performed with either passage 3 or 4 SARS-CoV-2. Infectious titers of SARS-CoV-2 were determined by plaque assay in Vero E6 cells in Minimum Essential Media supplemented with 4mM L-glutamine, 0.2% BSA, 10 mM HEPES, 0.12% NaHCO3, and 0.7% Oxoid agar (Cat #OXLP0028B). All SARS-CoV-2 infections were conducted in the CDC/USDA-approved BSL-3 facility of the Global Health and Emerging Pathogens Institute at the Icahn School of Medicine at Mount Sinai, in accordance with institutional biosafety requirements. Virus inoculation used a multiplicity of infection (MOI) of 0.1 or mock infection for 24 hours.

#### Compound screen

Imiquimod (Alfa Aesar, Cat # J63990MC), everolimus (Alfa Aesar, Cat # J60139MA), azathioprine (Alfa Aesar, Cat # J6231403), retinol (Alfa Aesar, Cat # J6207903), and the aHIV protease inhibitors; nelfinavir (MCE, Cat # HY-15287) and saquinavir (MCE, Cat # HY-17007) were reconstituted in 100% DMSO. Stocks were further diluted in DPBS to a working concentration of 100x. A549 cells expressing the ACE2 receptor were first treated with 10μM of each drug or 0.01% DMSO (negative control) for 24 hours in triplicate. We selected 10 μM as a commonly used screening concentration to detect strong inhibitory activity in cell-based assays.

Cells were then infected with SARS-CoV-2 at an MOI of 0.1 for 24 hours before harvesting cells in QIAzol lysis reagent.

#### SARS-CoV-2 quantification

RNA was isolated using the RNeasy mini kit (Qiagen, Cat # 74106) supplemented with RNAse inhibitor (Takara, Cat # 2313A) at 5% by volume. After quantification by QuBit, 400 ng of RNA was used to prepare cDNA via the High-Capacity cDNA Reverse Transcription Kit (Applied Biosystems, Cat # 4368813). TaqMan probes for the SARS-CoV-2 spike protein and control genes (GAPDH and Actin-B) were obtained from Thermo Fisher (Cat # 4331182). Real-time PCR was performed in triplicate using the Applied Biosystems QuantStudio 5. SARS-CoV-2 was quantified using the delta-delta Ct (2^−ΔΔCt^) method.

## Results

### Overview of the approach

Towards prioritizing readily available candidate compounds for potential preclinical or clinical testing (Fig. 1), we performed *in silico* computational drug repurposing (CDR) by integrating [6] genetically regulated gene expression (GReX) for COVID-19 associated hospitalization [9] (covering 17 tissues with significant gene-trait associations, including lung and blood; Table S1) [8] with a perturbagen signature library that includes readily repurposable medications [13]. This approach prioritizes compounds with the potential to reverse GReX profiles linked to COVID-19 susceptibility. To maximize translational potential, we restricted candidates to FDA-approved medications prescribed to individuals who tested positive for SARS-CoV-2 in the VHA. We subsequently *(1)* examined whether these candidates were associated with a decreased likelihood of testing positive for SARS-CoV-2, and *(2)* evaluated each compound’s *in vitro* activity against SARS-CoV-2 replication.

**Fig. 1.**
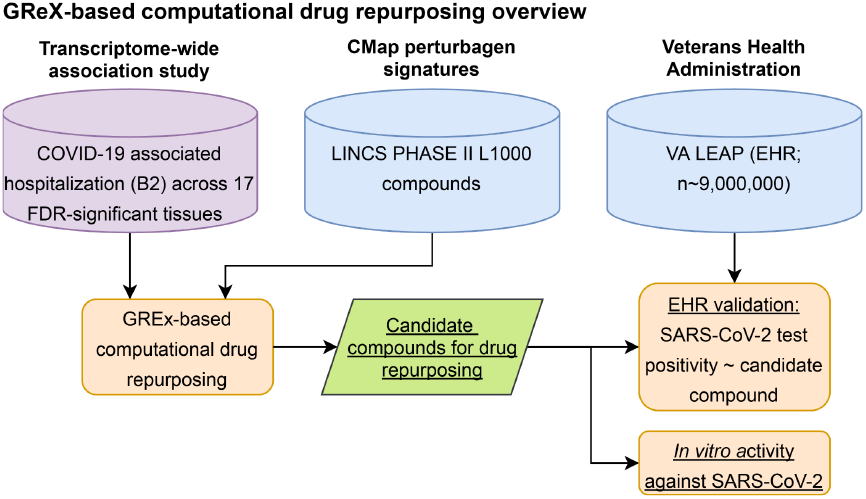
Data-driven GReX-based approach for computational drug repurposing in COVID-19. The schematic illustrates how genetically regulated gene expression (GReX) was combined with perturbagen signature libraries (LINCS) to identify candidate medications that may reverse the molecular dysregulation associated with COVID-19 susceptibility. Ranked compounds were subjected to two independent validation steps: *(1)* electronic medical records (EMR) validation based on the VHA’s Leveraging Electronic Health Information to Advance Precision Medicine (LEAP) program and *(2) in vitro* validation in an anti-SARS-CoV-2 replication assay in A549 cells.

### Genetically regulated gene expression (GReX-) based computational drug repurposing (CDR) analysis for COVID-19

The CDR pipeline ranked compounds based on their ability to antagonize the polygenic COVID-19 GReX signature, quantified by a pseudo-z-score (Table S2). We further limited selection to those with ranks significantly lower than the median (FDR-adjusted Mann-Whitney U test p < 0.05; Table S1). From this process, the top 10 candidates included imiquimod, nelfinavir, saquinavir, everolimus, azathioprine, nisoldipine, cerulenin, pyrvinium-pamoate, retinol, and selamectin (Table 1). The leading mechanism of action (MOA) was anti-HIV protease inhibition, with nelfinavir and saquinavir both appearing in the top 10 (Table 1).

**Table 1.**
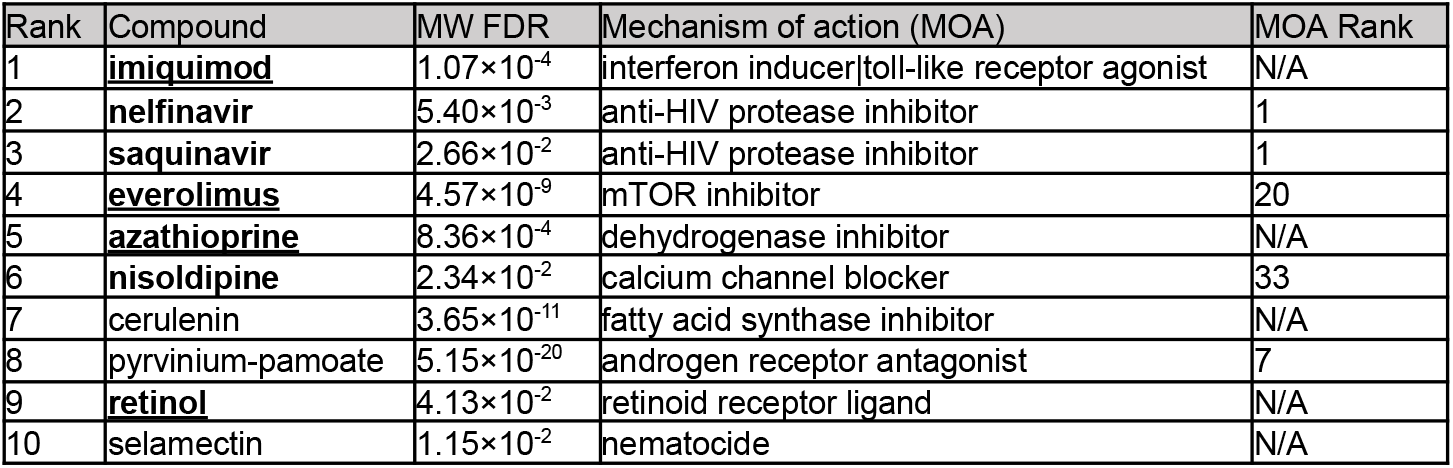
Top 10 candidate compounds identified by the GReX-based computational drug repurposing pipeline. FDA-approved compounds are indicated in bold, and compounds that were prescribed to at least 100 veterans tested for SARS-CoV-2 in the Veterans Health Administration (VHA) are underlined. The leading mechanism of action (MOA) is anti-HIV protease inhibition. Each compound’s “MW FDR” represents the Mann–Whitney U (MW) test p-values (estimated across all signatures to get the null distribution), adjusted for multiple testing (FDR; among considered “launched” compounds). “MOA Rank” refers to the rank of the mechanism of action among “launched” compounds.

| Rank | Compound | MW FDR | Mechanism of action (MOA) | MOA Rank |
| --- | --- | --- | --- | --- |
| 1 | <u>imiquimod</u> | $1.07 \times 10^{-4}$ | interferon inducer\|toll-like receptor agonist | N/A |
| 2 | **nelfinavir** | $5.40 \times 10^{-3}$ | anti-HIV protease inhibitor | 1 |
| 3 | **saquinavir** | $2.66 \times 10^{-2}$ | anti-HIV protease inhibitor | 1 |
| 4 | <u>everolimus</u> | $4.57 \times 10^{-9}$ | mTOR inhibitor | 20 |
| 5 | <u>azathioprine</u> | $8.36 \times 10^{-4}$ | dehydrogenase inhibitor | N/A |
| 6 | **nisoldipine** | $2.34 \times 10^{-2}$ | calcium channel blocker | 33 |
| 7 | cerulenin | $3.65 \times 10^{-11}$ | fatty acid synthase inhibitor | N/A |
| 8 | pyrvinium-pamoate | $5.15 \times 10^{-20}$ | androgen receptor antagonist | 7 |
| 9 | **retinol** | $4.13 \times 10^{-2}$ | retinoid receptor ligand | N/A |
| 10 | selamectin | $1.15 \times 10^{-2}$ | nematocide | N/A |

### Population-level and *in vitro* validation of top candidate compounds against COVID-19 incidence

After excluding cerulenin, pyrvinium-pamoate, and selamectin (none currently FDA-approved), we investigated whether any remaining candidates were associated with reduced COVID-19 incidence. From the broader VHA cohort of over 9 million U.S. veterans, 755,346 individuals received a SARS-CoV-2 test. We estimated the odds of a positive SARS-CoV-2 test if a candidate compound was prescribed within 90 days prior to testing. Because nelfinavir, saquinavir, and nisoldipine were each prescribed to fewer than 100 individuals (Table S3), they were excluded from the final population-level analysis, leaving imiquimod, everolimus, azathioprine, and retinol (Table 1). We also examined compounds by MOA; while no mechanism of action was significant when adjusting for multiple test corrections (Table S2), the top class was anti-HIV protease inhibitors (Table 1). Since there is evidence that some protease inhibitors developed against HIV can bind the 3C-like protease (3CL^pro^) of SARS-CoV-2 [19], an additional analysis was performed for anti-HIV protease inhibitors.

Among the four compounds tested individually (Table S9), azathioprine and retinol were each significantly associated with lower odds of testing positive for SARS-CoV-2 (Fig. 2A), after adjusting for important epidemiologic factors, medication indications, and propensity to be tested (Table S11). Adjusting for specific medication indications (Table S8) was critical, as it additionally controls for potential effects of the disease under treatment on COVID-19 incidence and disease-specific behavioral modification (e.g. immunocompromised patients may be more careful in adhering to rules or may get tested more frequently). Azathioprine (odds ratio=0.69, 95% CI: 0.62-0.77) and orally administered retinol (odds ratio = 0.81, 95% CI: 0.72-0.92) both exhibited a significant association with reduced COVID-19 incidence (Fig. 2A). Imiquimod—despite documented systemic absorption when applied topically in humans [20] and mouse models [21]—did not show a significant association, potentially due to insufficient blood levels for systemic effects. Everolimus also lacked significance, likely because of a smaller exposed population (an order of magnitude fewer individuals) and insufficient power.

**Fig. 2.**
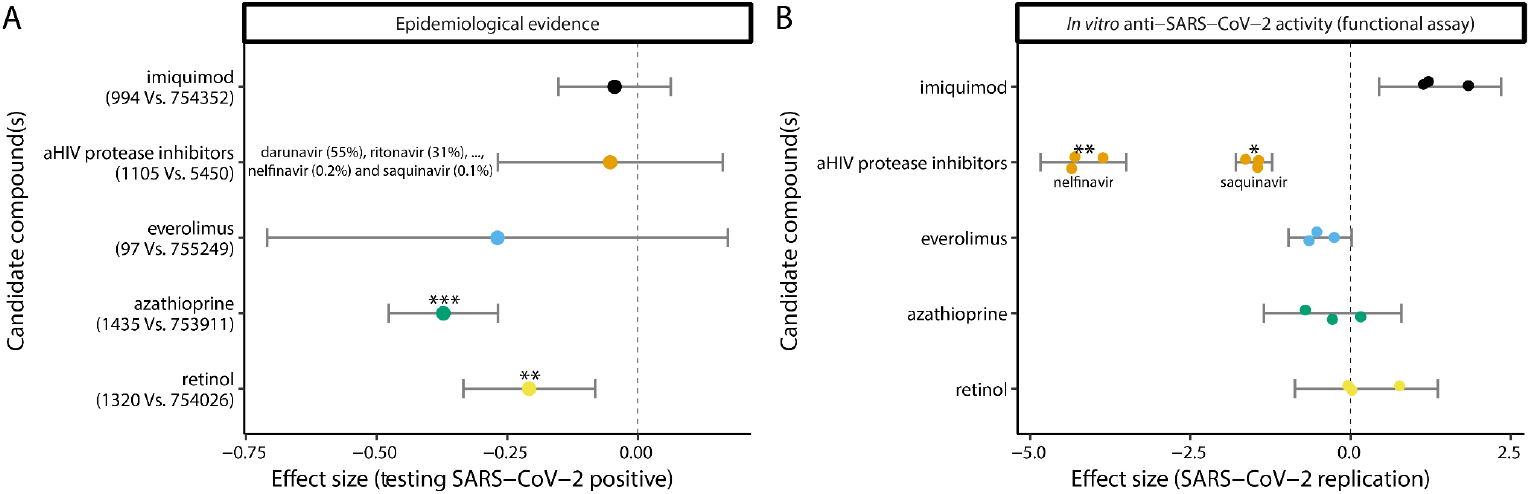
Epidemiological and *in vitro* validation of top candidates. **A**. Four individual compounds (imiquimod, everolimus, azathioprine, and retinol) and the highest-ranked medication category (anti-HIV protease inhibitors) were tested for their association with a reduced likelihood of a positive SARS-CoV-2 test. In total, 755,346 Veterans were included, and labels on the y-axis indicate the number of individuals who tested positive or negative. For the anti-HIV protease inhibitor category, participants receiving antiretroviral therapy with an anti-HIV protease inhibitor (n=1,105) were compared to those receiving other anti-HIV medications (n=5,450); the relative prescription frequency of anti-HIV protease inhibitors is also provided. The effect size (log(OR)) is plotted with 95% confidence interval (CI) error bars. ^***, **^, and ^*^ correspond to Bonferroni-adjusted p-values ≤0.001, ≤0.01, and ≤0.05, respectively. **B**. *In vitro* anti-SARS-CoV-2 activity was evaluated in ACE2-expressing A549 cells for imiquimod, nelfinavir, saquinavir, everolimus, azathioprine, and retinol (all at 10 μM for 24 hours). The effect on SARS-CoV-2 replication is shown as 2^−ΔΔCt^ (normalized to 0.01% DMSO vehicle) with 95% CI error bars (n=3). Negative values reflect reduced viral replication. ^***, **^, and ^*^ correspond to Bonferroni-adjusted p-values ≤0.001, ≤0.01, and ≤0.05, respectively.

For the leading MOA category (anti-HIV protease inhibitors), we analyzed HIV-positive patients on antiretroviral therapy, comparing those receiving protease inhibitors versus other antiretroviral regimens (Table S5). Despite similar comorbidity profiles between the two groups (Table S5), we detected no significant protective effect against SARS-CoV-2 (Fig. 2A; Table S10).

Following these epidemiologic analyses, we assessed the *in vitro* anti-SARS-CoV-2 activity of imiquimod, everolimus, azathioprine, retinol, nelfinavir, and saquinavir using A549 cells (adenocarcinomic human alveolar basal epithelial cells) expressing ACE2. Nelfinavir and saquinavir—ranked second and third, respectively (Table 1)—demonstrated robust anti-SARS-CoV-2 activity, with viral load reductions of approximately 95% and 65% (2^−ΔΔCt^ method), respectively (Fig. 2B). These results corroborate earlier findings in Vero E6 cells [22–24], underscoring the potential therapeutic relevance of these anti-HIV protease inhibitors.

## Discussion

Our study underscores the transformative potential of CDR as a strategy to accelerate the identification and validation of candidate therapeutics. By integrating GReX and perturbagen signature libraries (Fig. 1), we identified existing compounds that could be repositioned for COVID-19 [6, 7]. Previously, we validated a related pipeline by demonstrating that predicted therapeutics were progressively enriched for higher physician-curated indication levels across diverse disease categories (e.g., cardiovascular, autoimmune, neuropsychiatric) [7]. In this work, we employed an improved computational framework—a 5-method-rank approach [6] coupled with non-parametric aggregation and prioritization [8]—and extended its validation via *(1)* population-level analysis to assess whether predicted compounds were associated with reduced COVID-19 susceptibility, and *(2) in vitro* assays to evaluate anti-SARS-CoV-2 effects.

The strength of the CDR pipeline lies in its multi-tiered validation strategy. Our method capitalizes on the polygenic nature of gene expression dysregulation related to COVID-19–associated hospitalization. Compounds are first selected through GReX and perturbagen signature analyses, which identify agents that antagonize disease-linked imputed transcriptomes. We then incorporate real-world data from the Veterans Health Administration to evaluate whether these agents confer protective effects at the population level. Finally, we conduct *in vitro* assays to confirm their ability to inhibit SARS-CoV-2 replication. This layered approach increases confidence in candidates before dedicating the resources necessary for extensive clinical testing. Indeed, we identified multiple immunomodulators and two anti-HIV protease inhibitors as top candidates. Notably, azathioprine and retinol emerged as significantly protective against a positive SARS-CoV-2 test in our retrospective epidemiologic analysis (Fig. 2a), while nelfinavir and saquinavir demonstrated robust (~95% and ~65%, respectively) *in vitro* viral load reductions (Fig. 2b).

Although the pipeline was applied to COVID-19 as a proof of concept, its utility extends far beyond this specific context. The dual success of azathioprine and retinol in epidemiologic analyses underscores the promise of targeting immunomodulation in other conditions where immune system overactivation contributes to increased morbidity and mortality [25]. In parallel, nelfinavir and saquinavir’s strong *in vitro* anti-SARS-CoV-2 activity—despite low prescription rates (<0.5%) in the VHA population—highlights the potential for reusing less prescribed, often less-specific drugs. While initially focused on COVID-19, the core principle of harnessing host vulnerabilities via GReX-based computations is widely generalizable, offering a blueprint for rapid response in future outbreaks or for diseases with limited therapeutic options.

One of the key findings of this study is the importance of complementary validation approaches. In some cases (e.g., azathioprine or retinol), *in vitro* experiments using a simple alveolar epithelial cell system may fail to capture the immunomodulatory mechanisms that likely contribute to their protective signals at the population level [26–32]. Conversely, nelfinavir and saquinavir exhibited potent antiviral effects *in vitro*; however, their medication class showed no overall protective effect in the cohort, partly because the class was represented by other drugs with higher prescription rates including anti-HIV protease inhibitors that had previously been proven ineffective in clinical trials [33]. This underscores why a multilayered —computational, real-world epidemiological, and mechanistic—strategy is essential for prioritizing the most promising leads, while acknowledging that different validation modalities capture different biological dimensions.

Despite its advantages, the CDR pipeline is not without limitations. Our population-level analysis focused on COVID-19 incidence rather than disease severity, which was precluded by limited statistical power. As a result, we cannot infer whether azathioprine or retinol might reduce hospitalization or mortality. Additionally, among the seven FDA-approved compounds we investigated, only four were prescribed to at least 1,000 individuals in the 90-day window—an underrepresentation in a cohort of approximately seven million. Such constraints reflect the complexities inherent in real-world data, including prescription patterns and testing availability. Furthermore, the *in vitro* cell culture model does not recapitulate the complexities of *in vivo* immune interactions, which likely explains why certain immunomodulatory drugs were not similarly protective in the assay.

In conclusion, our CDR pipeline is a cost-effective and scalable approach to rapidly identify and validate candidate therapeutics. By uniting computational predictions, EMR-based population-level insights, and *in vitro* assays, we reduce the risk of pursuing false leads and clarify the mechanisms underlying observed clinical signals. Although demonstrated here in the context of COVID-19, the pipeline’s foundational principles—particularly its emphasis on targeting host vulnerabilities—can be extended to a wide array of complex diseases. Future studies integrating more extensive epidemiological data, preclinical animal models, and advanced molecular profiling techniques will help refine this approach and further expedite the discovery of high-impact therapies for diverse clinical challenges.

## Conclusions

Our study highlights the transformative potential of the CDR pipeline as a cutting-edge approach to accelerate the identification and validation of candidate therapeutics. To demonstrate its utility, we applied this pipeline to COVID-19, showcasing its ability to respond rapidly to a global health crisis requiring swift therapeutic discovery By integrating GReX-based pipelines with cost-effective pharmacoepidemiologic and *in vitro* analyses, we efficiently filtered a broad range of existing medications to prioritize those warranting deeper investigation which could provide further mechanistic insight. Among the six computationally predicted FDA-approved agents selected for validation (either prescribed to individuals tested for SARS-CoV-2 or belonging to the top-ranked class of anti-HIV protease inhibitors), four (66.67%; azathioprine, retinol, nelfinavir, and saquinavir) exhibited epidemiological or *in vitro* evidence of protective effects, though none exhibited both simultaneously. Future research, including large-scale clinical trials, mechanistic studies, and analyses in expanded population cohorts, will be essential to validate and refine candidate therapies.

## Data Availability

All data produced in the present work are contained in the manuscript.

## Data availability

All relevant information is included in the manuscript and supplementary tables.

## Abbreviations

COVID-19: Coronavirus Disease 2019
SARS-CoV-2: Severe Acute Respiratory Syndrome Coronavirus 2
GReX: Genetically Regulated Gene Expression
EMR: Electronic Medical Record
FDA: Food and Drug Administration
VHA: Veterans Health Administration
TWAS: Transcriptome-Wide Association Study
LINCS: Library of Integrated Network-Based Cellular Signatures OR Odds Ratio
CI: Confidence Interval
RT-PCR: Reverse Transcriptase Polymerase Chain Reaction
ACE2: Angiotensin-Converting Enzyme 2
BSL-3: Biosafety Level 3
MOI: Multiplicity of Infection
HIV: Human Immunodeficiency Virus
FDR: False Discovery Rate

## Supplementary Information

Additional File 1: Tables S1-11

## Author information

### Contributions

Conceptualization and study design: GV, JFF, JAL, PR. Data contribution or analysis tools: GV, KML, JMV, WZ, DH, SV, JB, MA, ZW, SR, LG, KC, JSL, SKI, S-WL, TLA, GEH, BRtO, JFF, JAL, PR. GV, KML, JMV, WZ, DH, SV, JB performed the analyses. GV, KML, JMV, JFF, JAL, PR wrote the manuscript with input from all authors.

## Ethics declarations

### Ethics approval and consent to participate

Analysis of national VA data was conducted under the protocol, “Leveraging Electronic Health Information to Advance Precision Medicine (LEAP)”, which was approved by VA Central Institutional Review Board and by the Research & Development Committees at Palo Alto, Salt Lake City, and West Haven VA Medical Centers.

### Consent for publication

Not applicable

